# Consent to Recontact for Future Research Using Linked Primary Healthcare Data: Outcomes and General Practice Perceptions from the ATHENA COVID-19 Study

**DOI:** 10.1101/2024.08.13.24311963

**Authors:** Kim Greaves, Amanda King, Zoltan Bourne, Jennifer Welsh, Mark Morgan, M. Ximena Tolosa, Trisha Johnston, Carissa Bonner, Tony Stanton, Rosemary Korda

## Abstract

**Background:** The ATHENA COVID-19 (ACV19) study was set up to recruit a cohort of patients with linked health information willing to be re-contacted in future to participate in clinical trials, and also to investigate the outcomes of people with COVID-19 in Queensland, Australia, using consent. This report describes how patients were recruited, their primary care data extracted, proportions consenting, outcomes of using the recontact method to recruit to a study, and experiences interacting with general practices requested to release the primary care data.

**Methods:** Patients diagnosed with COVID-19 from January 1^st^, 2020, to December 31^st^, 2020, were systematically approached to gain consent to have their primary healthcare data extracted from their general practice into a Queensland Health database and linked to other datasets for ethically approved research. Patients were also asked to consent to allow future recontact to discuss participation in clinical trials and other research studies. Patients who consented to recontact were later approached to recruit to a long-COVID study. Patients’ general practices were contacted to export the patient files. All patient and general practice interactions were recorded. Outcome measures were proportions of patients consenting to data extraction and research, permission to recontact, proportions of general practices agreeing to participate. A thematic analysis was conducted to assess attitudes regarding export of healthcare data, and the proportions consenting to participate in the long-COVID study also reported.

**Results:** Out of 1212 patients with COVID-19, contact details were available for 1155; 995(86%) were successfully approached, and 842(85%) reached a consent decision. Of those who reached a decision, 581(69%), 615(73%) and 629(75%) patients consented to data extraction, recontact, and both, respectively. 382 general practices were contacted, of whom 347(91%) had an electronic medical record compatible for file export. Of these, 335(88%) practices agreed to participate, and 12(3%) declined. In total 526 patient files were exported. The majority of general practices supported the study and accepted electronic patient consent as legitimate. For the long COVID study, 376(90%) of those patients recontacted agreed to have their contact details passed onto the long COVID study team and 192(53%) consented to take part in their study.

**Conclusions:** This report describes how primary care data was successfully extracted using consent, and that the majority of patients approached gave permission for their healthcare information to be used for research and be recontacted. The consent-to-recontact concept demonstrated its effectiveness to recruit to new research studies. The majority of general practices were willing to export identifiable patient healthcare data for linkage provided consent had been obtained.

## Background

Timely recruitment of suitable patients in sufficient numbers to clinical trials remains a major challenge to the research industry and the same applies to COVID-19 related research.^1^ Consent to recontact is a strategy whereby people give generic consent to be recontacted about new research opportunities.^2,3^ The availability of a registry of patients with linked health care information that can be screened to find specific patients for trials who have also pre-consented to be contacted to discuss participation in new research opportunities, would greatly accelerate clinical trial timelines and translation to outcomes.

The effective use of patient healthcare information in countries by linking primary care, hospital, and other health registry data to improve healthcare delivery is a global goal.^4^ Primary care data is particularly valuable as it contains detailed health information that is available at scale, with an estimated 80% of Australians visiting a general practitioner at least once each year.^5^ These linked data can be used for population health research, development of cohort studies and, if consent has been obtained, provide a large pool of participants that can be readily screened and then recontacted for recruitment to clinical trials.^6–9^ However, Australia has lagged behind other high-income nations in linking primary care data to other regional and national sources.^10^ Reasons for this include general practice clinics working as private companies, heterogenous software platforms that lack interoperability, limited incentives and concerns about sharing patient data.^11–15^ General practitioners are the custodians of primary care data in Australia and therefore it is essential to understand their attitudes towards sharing the data.^16^ Although a modest amount has been published regarding attitudes of general practitioners towards data sharing, little is known about their beliefs in Australia.^14,15, 17–21^

The ATHENA COVID-19 (ACV19) study was set up to recruit a cohort of patients with linked health information willing to be re-contacted in future to participate in clinical trials. It was also designed to enable ongoing investigation of health outcomes for people diagnosed with COVID-19 in Queensland, Australia, through the consent-based linkage of primary healthcare data with other datasets.^22^ The study was also designed to inform future implementation of the ATHENA program, a consent-based, state-wide, systematic mass recruitment of patients and linkage of healthcare data for the purposes of research and accelerated recruitment to clinical trials. This report describes how the study was set up, recruitment methods, and proportions consenting to primary care data linkage, and future recontact. The study also reports on the effectiveness of using the consent-to-recontact method to recruit to a new research study, and experiences interacting with general practices. The aim is to provide insights for those intending to use consent to extract, link and use primary healthcare data for research, and employ the recontact method to recruit to clinical trials.

## Methods

### Setting

The ATHENA program (Australians Together Health Initiative) is a Queensland Health funded concept involving the integration of primary, secondary and other healthcare datasets across Queensland. The vision is to deliver a state-wide registry containing the healthcare information and biospecimens of several million Australians across Queensland using dynamic consent to connect patients, researchers and the clinical trials industry.

As part of this, the ACV19 study was set up in 2020 in response to the COVID-19 pandemic to create a cohort of all people diagnosed with COVID-19 in Queensland with linked primary, secondary and registry data. There were two parts to the study. Part 1 (completed) linked Queensland COVID-19 hospital and administrative data including notifiable conditions and deaths, did not require informed consent and is described in detail elsewhere.^22^ Part 2 (described here) links data from the Part 1 study to patients’ healthcare information held within general practices and requires patient consent. All patients who had COVID-19 in Queensland in the period January 1^st^, 2020, up until December 31^st^, 2020, were contacted to gain consent to have their primary healthcare data extracted from their general practice into a Queensland Health database and linked to other datasets for ethically approved COVID-19 research. Patients were also asked to consent to be re-contacted about possible participation in future COVID-19 related studies.

In Australia, the vast majority of general practices use an electronic medical records system to keep coded records of patients.^23^ Multiple vendors exist of which Best Practice and Medical Director are the two most commonly used and have 90-95% of market share.^24^ The ACV19 study engaged with these two systems specifically because of their innate capacity to export data in XML format. Conversely, systems lacking this functionality, as well as practices unable to export data in XML format, were not eligible for inclusion in the study.’

### Patient recruitment

Patients identified as having had COVID-19 from the Queensland Health’s Notifiable Conditions System were telephoned by the ACV19 team to ask permission to email or post an information pack about the study. After 2-5 days the patient was recontacted to answer any further questions and guide them through the patient information consent form. Patients then electronically signed the form (DocuSign) or manually signed-and-returned it by post. Patients nominated up to a maximum of three general practices with whom they were now or previously registered. In Australia, a person can be registered with multiple practices. Patients were informed that their consent was being verbally recorded.

### General practice interaction

Prior to patient recruitment, the study was advertised in Primary Health Network newsletters and a website created containing study information, with supporting letters from the RACGP, Chief Health Officer and all seven Primary Health Networks in Queensland.^25^ An electronic clinical trials management system was used to manage and record all patient and general practice interactions. The liaison team consisted of a general practitioner (lead), two registered nurses, a Primary Health Network practice support officer and an allied health worker. An introductory call to the patient’s general practice checked practice details and confirmed they had one of the two electronic record systems suitable for data transfer. After five days, a second call confirmed information receipt, answered questions and confirmed the patient was registered with the practice. Here, it was explained their patient had consented to participate which required export of the identifiable whole patient file to the Queensland Health. It was explained that no financial reimbursement for data release was available.

### Data extraction from general practices

An email was sent to the practice manager’s inbox containing a single-use, secure file, transfer-hyperlink (Kiteworks) a standard method of file transfer used by Queensland Health and appended with the general practitioner’s provider number. No other patient or practitioner identifiers were contained in the message. A formal patient ‘records request’ letter and the patient’s completed patient information and consent form were also sent via secure electronic messaging (Medical Objects) to the nominated inbox within the general practice electronic medical record system. This contained documents pre-tagged with the patient’s name and date of birth to allow direct ingestion into the correct patient file within the system. Medical Objects is a widely used, secure method for information exchange between general practices and Queensland Health. A small minority of practices requested facsimile. The general practice exported the whole patient file to Queensland Health using Kiteworks. If no reply had been received within 5-7 days, the general practice was recontacted to guide them through the process. If the practice had any concerns, they were referred to the general practitioner lead who contacted the practice to resolve any issues. A small minority of practices requested facsimile. Contacting patients and general practices by the study team occurred between January 3^rd^, 2021 -August 27^th^, 2021. The ACV19 steering committee decided that once file transfer occurred and data was extracted, the file content would be deleted.

### Data linkage with Queensland health data

Primary healthcare data were linked probabilistically, using name, date of birth and address by the Statistical Services Branch, to Queensland COVID-19 hospital and administrative data including COVID-19 notifications and deaths from Part 1, applying established protocols.^26^

### Data analyses

General practice geographical area was classified using the Modified Monash Model which defines whether a location is a city, rural, remote or very remote (Modified Monash 1 is a major city, 7 very remote).^27^ To identify recurring responses and themes from interactions with the general practices, a thematic analysis was conducted by the project lead (KG) using a standard phased approach as a guide.^28^ Members of the general practice liaison team reviewed and familiarised themselves with the records of their general practice interactions and completed a questionnaire whereby they broadly listed practice types of responses. Based on these responses and known general practice attitudes to sharing data published in the literature, a list of categories was drawn up for focus group discussion by the project lead.^14, 15^ The discussion involved the whole team, was audio recorded, and conducted by the project lead, whereby each category and experience per team member was discussed in detail. The number of general practices who had responded in each category was recorded. The audio recording of the discussion was transcribed verbatim, and the transcripts reviewed over multiple inductive cycles from which broader general themes were developed.^28^

### Using the consent to recontact to recruit to a research study

A research group from University of Queensland studying long-COVID wished to use the recontact process to recruit additional participants from the ATHENA COVID-19 cohort into their ethically approved study. The research group had used social media to advertise nationally and recruited 80 patients. Details of their study can be found elsewhere.^29^ After protocol approval by the ACV19 steering committee, a proportion of the cohort who had consented to recontact were contacted and provided with an outline of the long-COVID study. Recontacted participants were asked if interested in hearing more about the study and, if so, gave consent to having their contact details passed onto the long COVID study team, who then contacted them to discuss the project in detail.

### Patient and public involvement

Between 2017-2019 a proof-of-concept study gauged the willingness of the public to link their healthcare data and received a positive response. One-on-one interviews were also undertaken with members of the public to understand the barriers and enablers to sharing and linking of primary healthcare data for research. The concept was also discussed with key stakeholders, including culturally and linguistically diverse community group leaders, and Australian and Torres Strait Islander representatives, to determine culturally sensitive approaches for recruitment of minority groups. This resulted in the Australian and Torres Strait Islander cohort recruitment being undertaken by a Queensland Health staff member identifying as Australian and Torres Strait Islander.

### Ethics approval

The ACV-19 study was granted ethics approval by the Gold Coast Human Research Ethics Committee 24^th^ April 2020, # HREC/2020/QGC/63555.

## Results

1212 patients were registered in the Notifiable Conditions System as having had COVID-19 from Jan 1^st^, 2020, to Dec 31^st^, 2020, in Queensland (Figure 1). Due to living interstate, 1155 participant contact details were listed to call, and 995(87%) were successfully contacted, of whom 842(85%) reached a consent decision. Of these, 581(69%) agreed to data extraction, 615(73%) to recontact, and 567(67%) to both (Table 1). Out of the 995 patients who were successfully contacted, 58%, 62% and 63% consented to data extraction, recontact, or both, respectively. The mean age of the cohort that reached a consent decision was 49.2 (95% range 21 - 77) years, and 50% were male. The mean age of those who consented was 50.6 (95% range 22 - 77) years and declined 46.1 (95% range 20 - 77) years. Patients were registered to 382 practices, of whom 347(91%) had a compatible electronic record system. 335(91%) practices agreed to transfer their files, 12 declined, 331(88%) practices successfully exported their data, and 4 practices were unable to transfer their files. A total of 520 files (497 individual patients) were exported; 21 patients had ≥2 records from multiple practices. The majority of practices had a single registered patient, most were located in cities and larger towns (Table 2).

**Figure 1.**
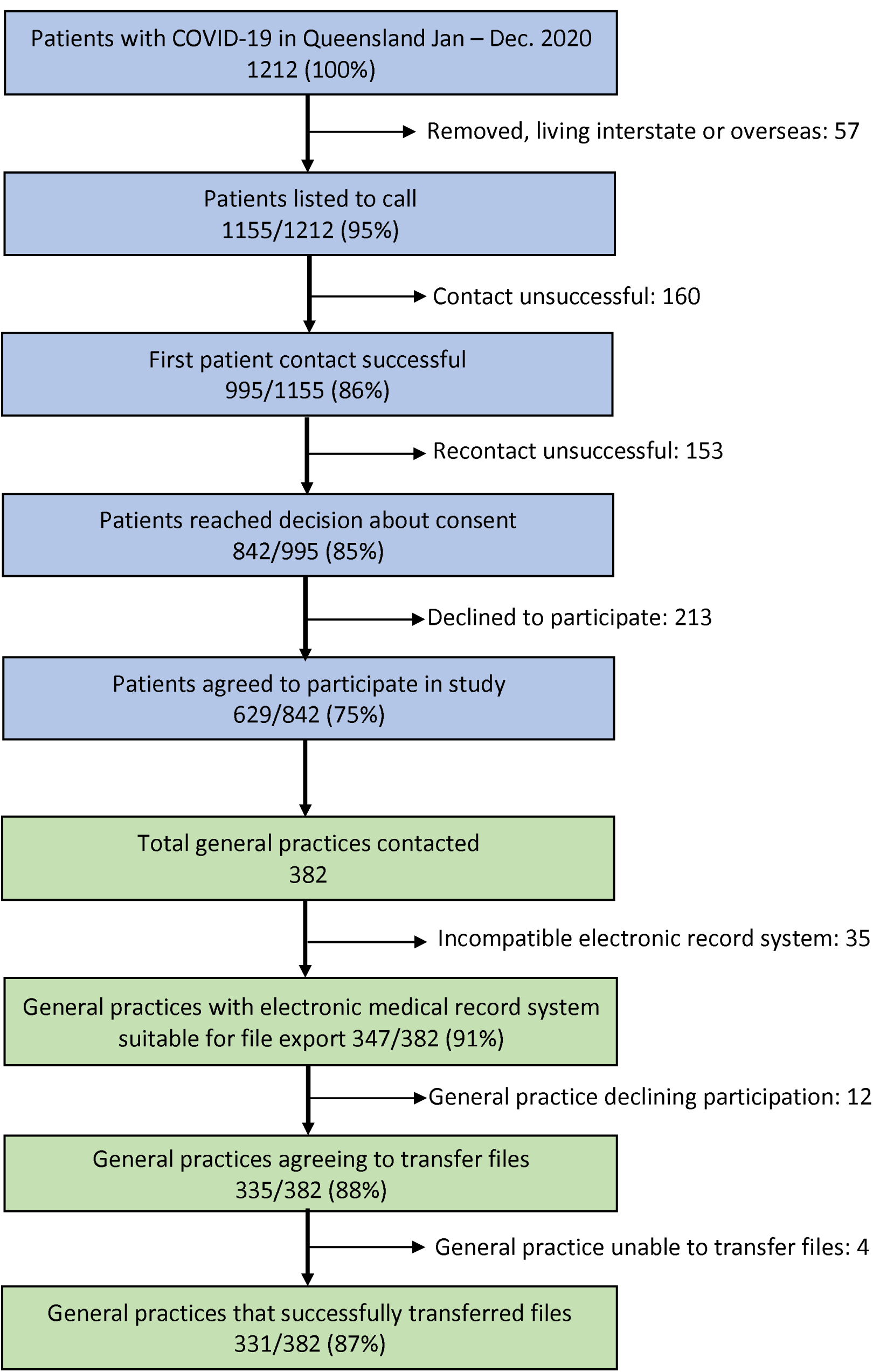
**Flow chart of patient and general practice recruitment**

**Table 1.** Categories of patient consent responses to request health data extraction from general practice and use for research, and recontact to discuss participation in future clinical trials.

|  | Consent to data extraction for research |  |  |  |
| --- | --- | --- | --- | --- |
| Consent to recontact |  | Agreed | Declined | Total |
|  | Agreed | 567(67%) | 48(6%) | 615(73%) |
|  | Declined | 14(2%) | 213(25%) | 227(27%) |
|  | Total | 581(69%) | 261(31%) | 842 |

**Table 2.**
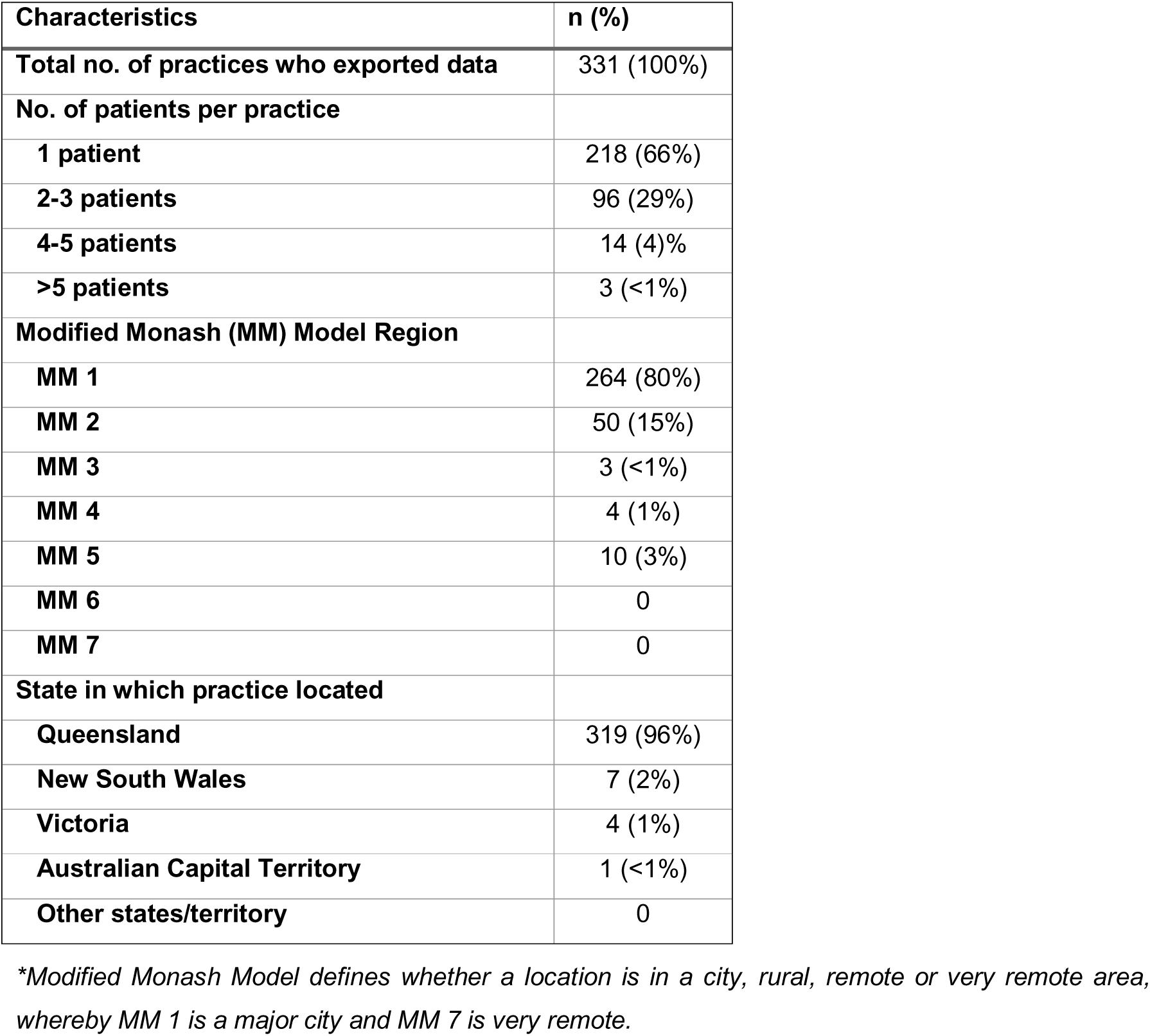
Characteristics of participating general practices and their patients with an electronic medical record-compatible export file.

### Consent to recontact to recruit to a research study

Of those patients who consented to recontact, 416 were contacted to discuss participation in the long-COVID study. Of these, 376(90%) consented to have their contact details passed onto the long-COVID team with 32(8%) declining, and 8(2%) were already enrolled. The long-COVID team approached 365 patients and 192(53%) consented to take part in their study.

### Interactions with general practice representatives grouped by theme

Interactions with general practices were grouped into five themes: 1) support for study concept and sharing of health information for research; 2) patient consent to allow healthcare data release; 3) trust, legal and cybersecurity issues; 4) reimbursement, time constraints and technology issues; 5) corporate-owned general practice. Details of the interactions are provided in Table 3 and brief outline is provided here.

**Table 3.**
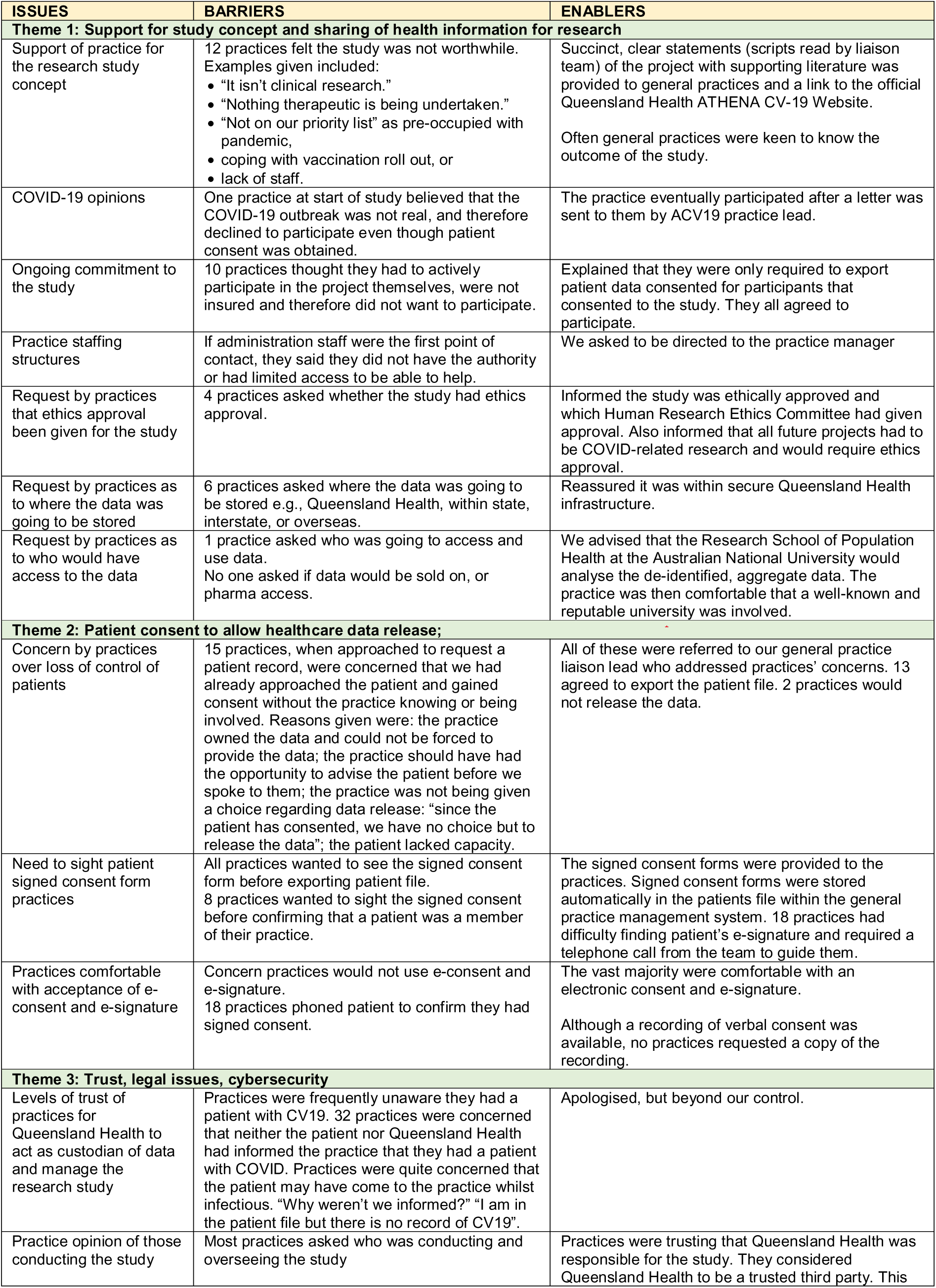
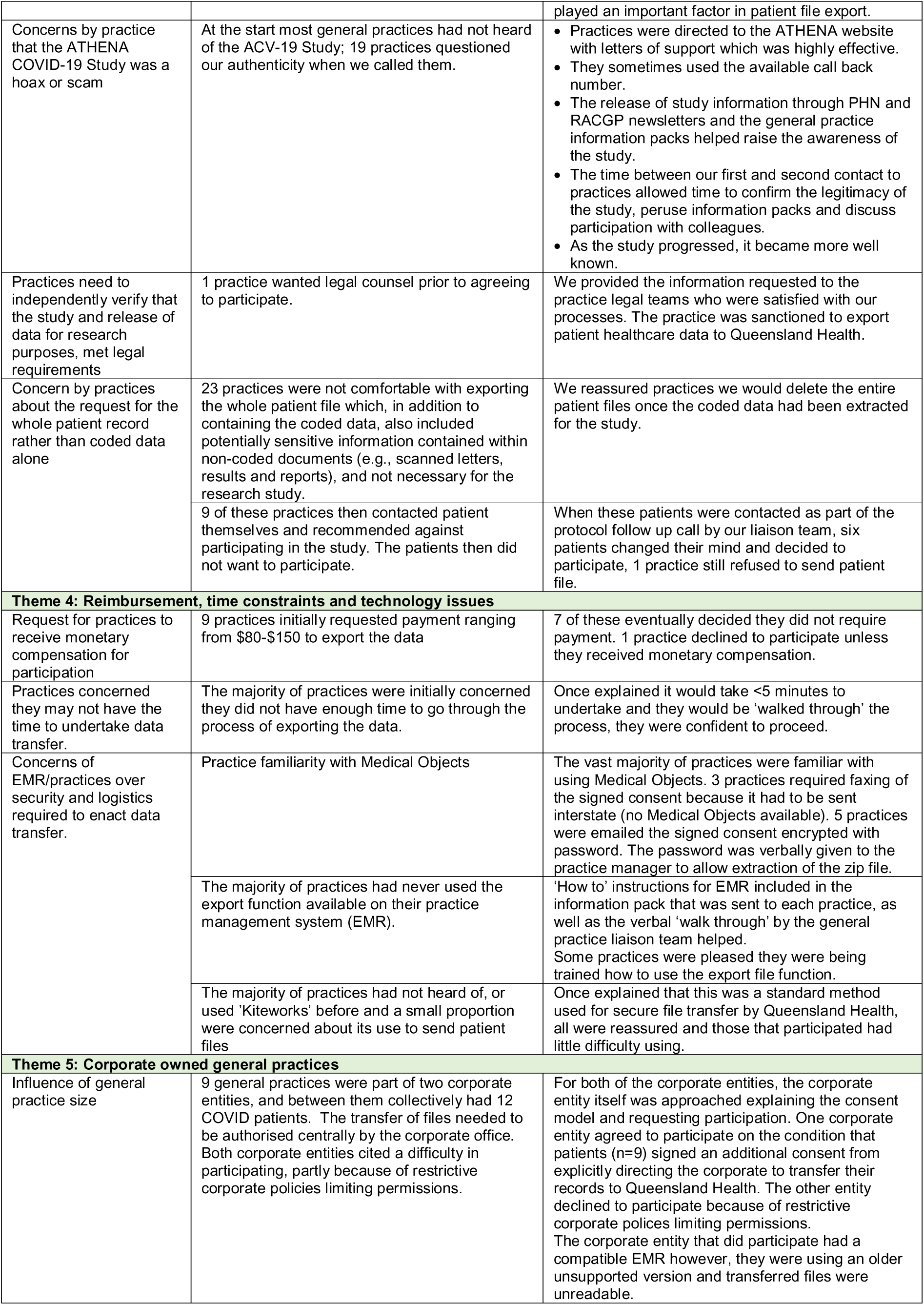
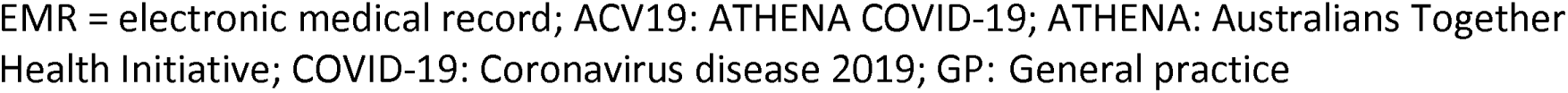
General practice attitudes to the export of primary healthcare data to Queensland Health for COVID-19 research.

### Theme 1. Support for study concept and sharing of health information for research

The majority response was “generally speaking, quite positive,” regarding “the idea of the study and contributing to the knowledge of COVID.” Twelve practices felt the study was not worthwhile. Ten thought they had to actively participate with concerns about lack of research experience and insurance cover. Four practices asked about ethics approval, and six about healthcare information storage (interstate or internationally) and all eventually took part. One practice wanted to know who had access to and could use the data.

### Theme 2. Patient consent for healthcare data release

Fifteen practices were concerned we had approached the patient and gained consent without the practice knowing or being involved. The perception by the liaison team was that practices believed they should have had the opportunity to advise the patient before we spoke to them. All practices made the eventual decision to release the data but two required further discussion with our general practitioner lead. All practices wanted a copy of the signed consent form before patient file export. The vast majority were comfortable with accepting the electronic consent form and electronic signature as a legitimate form of consent. Several general practices that did not accept electronic consent phoned the patient to confirm that they had signed the consent form.

### Theme 3. Trust, legal issues, cybersecurity

The majority of general practices had a high level of trust for Queensland Health to act as custodian of data and manage the research study. One team member stated: “I think it was a key factor that it was a government-led initiative.” Most practices at the start of the study had not heard of the ACV19 project. Nineteen practices thought the team caller was a scam when first contacted. This was managed by directing the general practice staff to the Queensland Health ACV19 website. One practice sought independent legal counsel about participation in the study and then took part. Some practices (n=23) had concerns about exporting the whole patient file to the Queensland Health database as this included non-coded documents such as scanned letters and imaging reports. Once made clear that the whole file would be deleted once the coded information has been extracted, the practices were comfortable with exporting files. Nine practices contacted the patients and recommended against participating. When these patients were contacted as part of the study follow up protocol, six patients changed their mind and participated.

### Theme 4. Reimbursement, time constraints and technology issues

Nine practices initially requested payment ranging from $80-$150 to export the data, however eight practices subsequently decided they did not require payment. The team reported that most practices were initially concerned they did not have enough time to export the data: However, once explained that the project team would assist, they agreed to proceed. From a technology perspective, although most practices were familiar with using Medical Objects, most had never used the export file function available in their electronic record system and required training. The majority of practices had not heard of Kiteworks and were concerned about using it. However, once explained it was a standard method used for file transfer by Queensland Health, practices were reassured.

### Theme 5. Corporate-owned general practices

Nine practices were part of corporate entities (two) with 12 patients collectively. File transfer needed authorisation, and only one corporate entity eventually allowed practice participation.

## Discussion

This study reports that the majority of patients gave permission for their healthcare information to be used for research – including that from primary care -and to be recontacted to discuss participation in clinical trials. The ATHENA consent-to-recontact concept also demonstrated its effectiveness to recruit to new research studies, with over 90% of patients recontacted agreeing to have their contact details passed on, and over 50% consenting to take part in a new research study. The study also describes the methods by which primary healthcare data can be successfully extracted from general practices and then linked for COVID-19 related research, using informed consent. Our study also reported the majority of general practices were willing to export identifiable patient healthcare data for linkage, provided consent had been obtained. Electronic consent was accepted by general practices as evidence of consent from patients. Lack of time and financial remuneration were not found to be significant issues. However, due to the relatively low consent proportions and risk of bias, investigation of the predictors of adverse outcomes was considered to lack validity and not worthwhile.

Slow recruitment of suitable participants to clinical trials is a major barrier facing research which delays the delivery of outcomes and inflates costs.^1^ Lower recruitment numbers also affect trial validity with little more than half of trials recruiting the originally specified sample size.^30, 31^ Novel approaches to recruitment strategies have been recommended and the consent-to-recontact method has been shown to be successful in the UK NHS Research for the Future program. This took a passive opportunistic approach using multi-media advertising to recruit patients.^2^ In contrast, our study took a systematic approach by actively contacting patients with COVID-19 and showed that the majority of patients approached were willing to be recontacted. Importantly, when applied to a real-world study, the recontact method successfully increased the number of patients recruited by 2.4-fold. Primary care in Australia has one of the most widespread uptake of electronic medical record systems in the world.^23^ Despite this, utilisation of primary healthcare data for research and other secondary purposes is not occurring.^32, 33^ Progress has been made with the introduction of the national ‘My Health Record’ and New South Wales ‘Lumos’ program. Lumos has linked the primary healthcare records of 1.3 million patients from 156 practices within New South Wales to other health system data.^34, 35^ However, due to privacy issues related to either an opt-out process (My Health Record) and a waiver of consent (Lumos), access to these datasets are restricted and use for research and consent to recontact is not possible.

The proportions consenting in our study were similar to other Australian studies.^36^ The relatively lower consent proportions (75%) suggest that a consent-based approach to gain permission to extract and link primary care data for population health research is not suitable due to the risk of bias. However, this approach is suitable for internal comparison research and mandatory for the creation of a cohort of patients who can be recontacted to participate in clinical trials. In this study, there are advantages and disadvantages to using either ‘patients reached a decision about consent’ (n=842), or where ‘first contact was successful’ (n=995), as a denominator, and therefore both values are quoted. Using the former as the denominator provides a focused analysis specifically on the subset of patients who actively participated in the consent decision process, and highlights consent rates among engaged participants. However, this method may also underestimate the overall participation rate whereby a significant number of patients who were successfully contacted did not reach a consent decision for various reasons (e.g., lack of interest, misunderstanding of study details). The advantage of the latter approach is that it offers a broader perspective on patient engagement regardless of the final consent decision.

In our study, patient consent was required as identifiable patient data was being extracted.^37^ A systematic review of 25 studies from multiple countries (including Australia) on the public responses to the sharing and linkage of health data for research purposes reported that there was widespread public support for research.^38^ However, support was conditional and depended on the interplay of multiple factors. Patients wanted control of their data (through consent) and the ability to express preferences as to who accessed their data. They also required a high level of trust in the institution looking after their data and knowledge of how it was to be used. Patients were sensitive to the fact that the need for consent had to be balanced against the inefficiencies of having to gain consent, and that lower consent proportions might lead to bias. The report noted that if participants understood the reasons for requiring the data, then opinions shifted away from opt-in consent to either opt-out or varied consent.

Since general practices are integral to the sharing and linkage of primary healthcare data, an understanding of their perspectives is vital. General practitioner attitudes to data sharing for research in Europe and the USA are well described.^17–20, 39^ However, little is known in the Australian context whereby previous studies have interviewed small numbers of general practitioners on their perspectives of the sharing of data for research.^14, 15^ Our study differs in that we collated the impressions from over 300 general practices asked to participate in a research study requiring healthcare data export. General practices are custodians of their patients’ health information and highly protective of patients’ privacy.^40^ This was evident in our study, where all practices wanted a copy of the signed consent, with a proportion phoning the patient to confirm. Interestingly, no general practice asked whether the data would be sold on or whether the pharmaceutical industry would have access which were concerns found in other studies.^14, 15^ In addition, some practices challenged the need to export the whole patient file indicating an awareness that only data relevant to the research study should be collected. Concerns around data security is a recurring theme in the literature mainly regarding patient re-identification and data breaches, but this was not an issue for the majority of general practices in our study.^14, 15^ Trust in the institutions conducting the research is also an important issue raised by general practitioners.^14, 15^ In our study, the impression was that practices felt reassured that Queensland Health and a reputable university were undertaking the study. Financial reimbursement to general practices for the time and effort taken to share health information is a common theme raised in the literature. In one study, all general practitioners believed remuneration to be a key issue and in another, two out of 11 felt a monetary incentive should be provided.^14, 15^ In our study only one practice (<1%) decided not to participate due to lack of payment. At the start of the study, we were concerned how practices would respond to the use of electronic consent. We found the majority accepted this method. This was likely due to the change in health practices induced by the pandemic. Social distancing and a greater reliance on electronic means of recording transactions such as electronic prescribing, meant that practices were already well on the path to accepting e-consent and digital signatures.^41^

Based on our findings, our recommendations would be to consider electronic rather than paper-based consent, active engagement with patients and taking a systematic (rather than opportunistic) approach when mass-recruiting for potential research and recontact. It is also important to ensure clear and transparent communication with patients about their data use and future recontact, and the importance of patient consent and active engagement with general practices if primary care data is to be used.

### Study limitations

A main limitation is that questionnaires were not used to directly assess general practice staff attitudes to sharing primary care data. Rather, we recorded interactions with staff over the course of the study and reported on them. Although all interactions included written documentation of comments made by general practice staff, subjective impressions were made by the liaison team and at risk of interpretation bias. Previous studies focused on the general practitioner’s attitude alone and did not encompass the opinions of all staff working in the practice. This should be considered when comparisons are made. Our study was conducted during the unique situation of a pandemic, and it may be suggested that once normal conditions resume, the results of this study will no longer apply. However, whilst the pandemic has caused a major shift in work patterns, the IT methods used in this study were already in standard use, and any new adaptations such as e-consent, are likely to remain permanent. Finally, 30% of patients and their associated practices did not participate in the study, which may be a source of selection bias. Despite these limitations, this study was able to elicit useful information from Australian general practices regarding their impressions on the export of health data for research. The extraction of general practice patient data was a time-consuming process and not suitable to be undertaken at scale. At the start of the pandemic this process was felt to be the most straightforward to set up, and whilst it has provided valuable information on processes and attitudes of general practices involved, a faster automated version of primary care data extraction is in development.

### Conclusions

This study showed that the majority of patients and their general practices consented to share their health information for research and recontact. The consent-to-recontact method proved successful in recruiting to a separate research study. The study also described the methods by which primary healthcare data was successfully extracted from general practices and linked to other health data for the purposes of research using informed consent. This knowledge will inform implementation of the larger ATHENA study, as well as those intending to link primary healthcare data for research and use for recruitment to clinical trials and other studies in future.

## Data Availability

All data produced in the present study are available upon reasonable request to the authors

## List of Abbreviations

ACV19: ATHENA COVID-19
ATHENA: Australians Together Health Initiative
COVID-19: Coronavirus disease 2019
GP: General practice

## Trial registration details

not applicable

## Declarations

### Ethics approval and consent to participate

The ATHENA COVID-19 Study was granted Ethics approval from the Gold Coast Human Research Ethics Committee, Queensland, Australia on the 24th April 2020 and all participants provided written informed consent before participating in the study. The study was conducted in accordance with the 1964 Helsinki declaration and its later amendments. Approval #: HREC/2020/QGC/63555

### Consent for publication

Not applicable

### Availability of data and materials

Datasets used and analysed during this study are available from the corresponding author upon reasonable request.

### Authors’ contributions

The authors confirm contribution to the paper as follows: study conception and design: KG, AK, ZB, JW, MM, XT, TJ, CB, TS, and RK; data collection: KG, AK, ZB, XT; analysis and interpretation of results: KG, AK, ZB, JW, MM, XT, TJ, TS, and RK; draft manuscript preparation: KG, AK, ZB, JW, MM, XT, TJ, CB, TS, and RK; manuscript copy-editing: KG, AK, ZB, JW, MM, XT, TJ, CB, TS, and RK. All authors reviewed the results and approved the final version of the manuscript.

## Acknowledgements

We would like to thank the team at the Health Innovation Investment and Research Office for their invaluable support. Also, the support of the ATHENA Coordination Centre, general practice liaison team of, Nikitas Kamarinos, Lisa Edward, Clinton Bazely and Kimberley Getley, also, Dr Jonathan Harper and Pattie Hudson at the Central Queensland, Wide Bay and Sunshine Coast PHN and Consumer advocate Anna Voloschenko.

## Authors’ information (optional)

None

## References

1. Al-Shahi Salman R, Beller E, Kagan J, et al. Increasing value and reducing waste in biomedical research regulation and management. Lancet 2014; 383: 176–185. 2014/01/15. DOI: 10.1016/S0140-6736(13)62297-7.

2. Grady K, Gibson M and Bower P. Can a ’consent to contact’ community help research teams overcome barriers to recruitment? The development and impact of the ’Research for the Future’ community. BMC Med Res Methodol 2019; 19: 195. 2019/10/24. DOI: 10.1186/s12874-019-0843-4.

3. Grant A, Ure J, Nicolson DJ, et al. Acceptability and perceived barriers and facilitators to creating a national research register to enable ’direct to patient’ enrolment into research: the Scottish Health Research Register (SHARE). BMC Health Serv Res 2013; 13: 422. 20131018. DOI: 10.1186/1472-6963-13-422.

4. World Health Organisation. Stronger Collaboration, Better Health. Global Action Plan for Healthy Lives and Well-being for All, https://www.who.int/initiatives/sdg3-global-action-plan (2019, accessed 20 Oct 2021 2021).

5. Britt H, Miller G, Henderson J, et al. General practice activity in Australia 2015–16, https://ses.library.usyd.edu.au/handle/2123/15514 (2016, accessed 20 Oct 2021 2021).

6. Brown A, Kirichek O, Balkwill A, et al. Comparison of dementia recorded in routinely collected hospital admission data in England with dementia recorded in primary care. Emerg Themes Epidemiol 2016; 13: 11. 2016/11/02. DOI: 10.1186/s12982-016-0053-z.

7. Einarsdottir K, Preen DB, Emery JD and Holman CD. Regular primary care decreases the likelihood of mortality in older people with epilepsy. Med Care 2010; 48: 472–476. 2010/04/16. DOI: 10.1097/MLR.0b013e3181d68994.

8. Einarsdottir K, Preen DB, Sanfilippo FM, et al. Mortality in Western Australian seniors with chronic respiratory diseases: a cohort study. BMC Public Health 2010; 10: 385. 2010/07/02. DOI: 10.1186/1471-2458-10-385.

9. Herrett E, Shah AD, Boggon R, et al. Completeness and diagnostic validity of recording acute myocardial infarction events in primary care, hospital care, disease registry, and national mortality records: cohort study. BMJ 2013; 346: f2350. 2013/05/23. DOI: 10.1136/bmj.f2350.

10. Productivity Commission Canberra. Data Availability and Use: Overview & Recommendations, https://www.pc.gov.au/inquiries/completed/data-access/report/data-access-overview.pdf (2017, accessed 20 Oct 2021 2021).

11. Boyle DI. Middleware Supporting Next Generation Data Analytics in Australia. Stud Health Technol Inform 2015; 216: 1019. 2015/08/12.

12. Palamuthusingam D, Johnson DW, Hawley C, et al. Health data linkage research in Australia remains challenging. Intern Med J 2019; 49: 539–544. 2019/04/09. DOI: 10.1111/imj.14244.

13. The Royal Australian College of General Practitioners. RACGP Practice Technology and Management: Minimum requirements for general practice clinical information systems to improve usability, https://www.racgp.org.au/FSDEDEV/media/documents/Running%20a%20practice/Support%20and%20tools/Minimum-requirements-for-general-practice-CIS.pdf (2018, accessed 20 Oct 2021 2021).

14. Hodgkins AJ, Mullan J, Mayne DJ, et al. Australian general practitioners’ attitudes to the extraction of research data from electronic health records. Aust J Gen Pract 2020; 49: 145–150. 2020/03/03. DOI: 10.31128/AJGP-07-19-5024.

15. Monaghan T, Manski-Nankervis JA and Canaway R. Big data or big risk: general practitioner, practice nurse and practice manager attitudes to providing de-identified patient health data from electronic medical records to researchers. Aust J Prim Health 2020; 26: 466–471. 2020/12/10. DOI: 10.1071/PY20153.

16. Royal Australian College of General Practitioners. Guiding principles for managing requests for the secondary use of de-identified general practice data, https://www.racgp.org.au/running-a-practice/security/managing-practice-information/secondary-use-of-general-practice-data/de-identified-general-practice-data (2019, accessed 21 Sept 2022).

17. Gentil ML, Cuggia M, Fiquet L, et al. Factors influencing the development of primary care data collection projects from electronic health records: a systematic review of the literature. BMC Med Inform Decis Mak 2017; 17: 139. 2017/09/28. DOI: 10.1186/s12911-017-0538-x.

18. Hummers-Pradier E, Scheidt-Nave C, Martin H, et al. Simply no time? Barriers to GPs’ participation in primary health care research. Fam Pract 2008; 25: 105–112. 2008/04/18. DOI: 10.1093/fampra/cmn015.

19. Spencer K, Sanders C, Whitley EA, et al. Patient Perspectives on Sharing Anonymized Personal Health Data Using a Digital System for Dynamic Consent and Research Feedback: A Qualitative Study. J Med Internet Res 2016; 18: e66. 2016/04/17. DOI: 10.2196/jmir.5011.

20. Stevenson F. The use of electronic patient records for medical research: conflicts and contradictions. BMC Health Serv Res 2015; 15. DOI: 10.1186/s12913-015-0783-6.

21. Stevenson F, Lloyd N, Harrington L and Wallace P. Use of electronic patient records for research: views of patients and staff in general practice. Fam Pract 2013; 30: 227–232. 2012/11/08. DOI: 10.1093/fampra/cms069.

22. Welsh J, Korda RJ, Paige E, et al. The ATHENA COVID-19 Study: Cohort profile and first findings for people diagnosed with COVID-19 in Queensland, 1 January to 31 December 2020. Commun Dis Intell (2018) 2021; 45 2021/10/01. DOI: 10.33321/cdi.2021.45.51.

23. Oderkirk J. Readiness of electronic health record systems to contribute to national health information and research, https://www.oecd-ilibrary.org/social-issues-migration-health/readiness-of-electronic-health-record-systems-to-contribute-to-national-health-information-and-research_9e296bf3-en (2017, accessed 20 Oct 2021 2021).

24. Macdonald K. Silly sausages being touted for Medical Director, https://www.pulseitmagazine.com.au/blog/6107-silly-sausages-being-touted-for-medicaldirector (2021, accessed October 20 2021 2021).

25. Health Innovation Investment and Research Office and Queensland Health. ATHENA COVID-19 Study, https://www.health.qld.gov.au/research-reports/research-projects/athena-covid-19 (2020, accessed October 20 2021 2021).

26. Queensland Government Department of Health. Queensland Data Linkage Framework. In: Statistical Analysis and Linkage Unit SSB, Department of Health, (ed.). Brisbane: State of Queensland (Queensland Health), 2021.

27. Department of Health Australian Government. Modified Monash Model. The Modified Monash Model (MMM) defines whether a location is a city, rural, remote or very remote. Read about the MMM and how to search for areas classified under its structure, https://www.health.gov.au/health-topics/health-workforce/health-workforce-classifications/modified-monash-model (2021, accessed 20 Oct 2021 2021).

28. Lester J, Cho Y and Lochmiller C. Learning to do qualitative data analysis: a starting point. Human Resource Development Review 2020; 19: 94–106.

29. Sinclair JE, Vedelago C, Ryan FJ, et al. Cardiovascular symptoms of PASC are associated with trace-level cytokines that affect the function of human pluripotent stem cell derived cardiomyocytes. bioRxiv 2024: 2024.2004.2011.587623. DOI: 10.1101/2024.04.11.587623.

30. Campbell MK, Snowdon C, Francis D, et al. Recruitment to randomised trials: strategies for trial enrollment and participation study. The STEPS study. Health Technol Assess 2007; 11: iii, ix–105. 2007/11/15. DOI: 10.3310/hta11480.

31. Sully BG, Julious SA and Nicholl J. A reinvestigation of recruitment to randomised, controlled, multicenter trials: a review of trials funded by two UK funding agencies. Trials 2013; 14: 166. 2013/06/14. DOI: 10.1186/1745-6215-14-166.

32. Sturgiss E and van Boven K. Datasets collected in general practice: an international comparison using the example of obesity. Aust Health Rev 2018; 42: 563–567. 2018/06/05. DOI: 10.1071/AH17157.

33. Tran B, Straka P, Falster MO, et al. Overcoming the data drought: exploring general practice in Australia by network analysis of big data. Med J Aust 2018; 209: 68–73. 2018/07/07. DOI: 10.5694/mja17.01236.

34. Australian Digital Health Agency. Benefits of My Health Record for healthcare professionals, https://www.myhealthrecord.gov.au/for-healthcare-professionals/what-is-my-health-record/benefits-my-health-record-for-healthcare (2021, accessed 20 Oct 2021 2021).

35. Correll P, Feyer A, Phan P, et al. Lumos: a statewide linkage programme in Australia integrating general practice data to guide system redesign. Integrated Healthcare Journal 2021; 3: e000074. DOI: 10.1136/ihj-2021-000074.

36. Graves A, McLaughlin D, Leung J and Powers J. Consent to data linkage in a large online epidemiological survey of 18-23 year old Australian women in 2012-13. BMC Med Res Methodol 2019; 19: 235. 2019/12/13. DOI: 10.1186/s12874-019-0880-z.

37. Australian Government and Office of the Information Commissioner. Privacy Act 1988. Federal Register of Legislation 1988.

38. Aitken M, de St Jorre J, Pagliari C, et al. Public responses to the sharing and linkage of health data for research purposes: a systematic review and thematic synthesis of qualitative studies. BMC Med Ethics 2016; 17: 73. 2016/11/12. DOI: 10.1186/s12910-016-0153-x.

39. Wyatt D, Cook J and McKevitt C. Perceptions of the uses of routine general practice data beyond individual care in England: a qualitative study. BMJ Open 2018; 8: e019378. 2018/01/11. DOI: 10.1136/bmjopen-2017-019378.

40. The Royal Australian College of General Practitioners. Secondary use of general practice data, https://www.racgp.org.au/download/Documents/e-health/Secondary-use-of-general-practice-data.pdf (2017, accessed 20 Oct 2021 2021).

41. Department of Health Australian Government. Providing health care remotely during COVID-19, https://www.health.gov.au/news/health-alerts/novel-coronavirus-2019-ncov-health-alert/coronavirus-covid-19-advice-for-the-health-and-disability-sector/providing-health-care-remotely-during-covid-19 (2021, accessed 20 Oct 2021 2021).

